# Structural Evaluation of *RYR2*-CPVT Missense Variants and Continuous Bayesian Estimates of their Penetrance

**DOI:** 10.1101/2025.03.20.25324327

**Authors:** Kohei Yamauchi, Matthew Ku, Devyn W. Mitchell, Alex Shen, Kundivy Dauda, Loren Vanags, Jeffrey Schmeckpeper, Bjorn C. Knollmann, Matthew J. O’Neill, Brett M. Kroncke

**Affiliations:** Department of Cardiovascular Medicine, Shiga University of Medical Science, Otsu, Shiga Prefecture, Japan; Division of Clinical Pharmacology, Department of Medicine, Vanderbilt University Medical Center, Nashville, TN, United States; Division of Cardiology, Department of Medicine, Vanderbilt University Medical Center, Nashville, TN, United States; Department of Medicine, Brigham and Women’s Hospital, Boston, MA, United States; Clinical Fellow, Harvard Medical School, Boston, MA, United States

**Keywords:** Penetrance, Catecholaminergic Polymorphic Ventricular Tachycardia, Arrhythmias, Statistical Genetics, Electrophysiology

## Abstract

**Background:** Catecholaminergic Polymorphic Ventricular Tachycardia (CPVT) is strongly associated with rare missense variants in *RYR2*, the gene encoding the intracellular calcium release channel RyR2. Precision medicine is complicated by incomplete penetrance, particularly in the case of *RYR2*-CPVT variants

**Objective:** To improve structural understanding and clinical actionability of *RYR2-*CPVT incomplete penetrance.

**Methods:** We curated 179 manuscripts reviewed by three individuals to extrapolate *RYR2*-CPVT missense variant genotype-phenotype relationships. Purportedly neutral control variants were ascertained from *RYR2* missense variants observed in gnomAD and ClinVar. We performed an *RYR2*-CPVT Bayesian penetrance analysis by conditioning a CPVT penetrance prior on variant-specific features (*in silico* and structural) calibrated by heterozygote phenotypes. We compared the calibration of our Bayesian penetrance estimates and our previous described structural density metric with *in silico* predictors REVEL, AlphaMissense and ClinVar annotations, using Spearman rank-order correlations, and Brier Scores. Penetrance estimates were superimposed upon a cryo-EM structure of RyR2 to investigate ‘hot-spot’ heterogeneity.

**Results:** From the literature and gnomAD, we identified 1,014 affected missense *RYR2* heterozygotes (468 unique variants) among a total of 622,575 heterozygotes (5,181 unique variants). Among the predictors, our Bayesian prior score had the highest Spearman rank-order and lowest Brier scores, respectively (0.19; 0.0090), compared to ClinVar (0.083; 0.019), REVEL (0.16; 0.018), or AlphaMissense (0.18; 0.018). Penetrance estimates for all *RYR2* missense variants are prospectively hosted at our Variant Browser website.

**Conclusions:** Bayesian penetrance scores outperform current tools in evaluating variant penetrance. We provide prospective CPVT penetrance values for 29,242 *RYR2* missense variants at our online Variant Browser.

## Introduction

Inherited arrhythmia syndromes commonly arise from variants in genes encoding cardiac ion channels, their modulating proteins, and cardiomyocyte structural proteins^1–3^. Catecholaminergic polymorphic ventricular tachycardia (CPVT) is one such rare syndrome resulting in atrial and ventricular arrhythmias, most frequently during periods of high adrenergic stimulus. Studies of the genetic basis of CPVT have revealed both autosomal dominant and autosomal recessive patterns of inheritance. Recently, the ClinGen variant curation working group identified strong evidence for the involvement of the following genes in CPVT: *RYR2, CASQ2, TECRL, TRDN,* and *CALM1-3* (Figure 1A)^3^. *RYR2* encodes for a calcium-activated calcium-release channel located intracellularly in the sarcoplasmic reticulum (SR), the major Ca*^2+^* storage organelle in cardiac muscle. During excitation-contraction coupling, RyR2 is activated by Ca^2+^ influx from voltage-gated L-type Ca^2+^ channels located in the cell membrane, a process known as Ca-induced Ca-release. Autosomal dominant RYR2 gain-of-function variants predispose to spontaneous channel openings and SR Ca^2+^ release during diastole, resulting in ventricular ectopy and triggers of re-entrant ventricular arrhythmias. Unfortunately, these events can result in sudden cardiac death (SCD) and may be the first manifestation of an otherwise unrecognized disease. The vast majority of CPVT cases associated with gain-of-function variants in *RYR2* compared to all other CPVT-linked disease genes shown in Figure 1A^3^.

**Figure 1.**
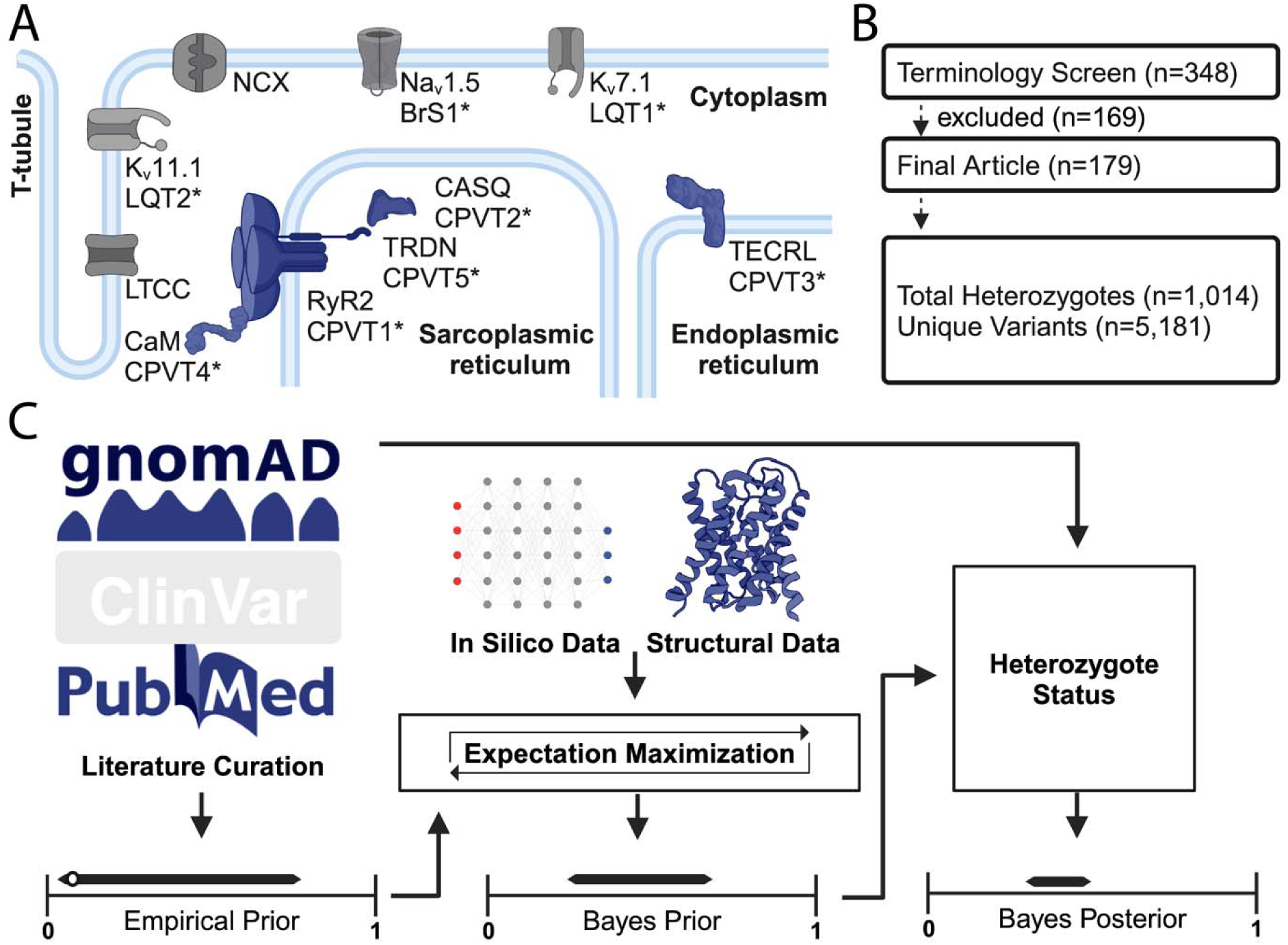
Overview Schematic of CPVT-associated proteins. A. Cellular schematic depicting key cardiomyocyte ion channels. These include ion channels with associated with other ion channelopathies, Nav1.5, Kv11.1, and Kv7.1 (Brugada Syndrome, Long QT syndrome type 2, and Long QT syndrome type 1, respectively), as well as key proteins associated with CPVT subtypes (RyR2, CASQ, TECRL, TRDN, and CaM). B. Flowchart depicting literature curation. C. Graphical schematic of Bayesian penetrance model.

Early identification of *RYR2*-CPVT variant heterozygotes may help prevent SCD if appropriate treatment is initiated. Traditionally, this has been done by family cascade screening of individuals with cardiac events later found to harbor deleterious *RYR2* variants. A complementary and emerging strategy is to use genotype-first approaches to pre-emptively identify genetically at-risk individuals before the emergence of events^4,5^. Progress towards this goal has been stymied by the frequency of genetic noise occurring in this large protein (15 kb), the lack of high-throughput functional approaches to characterize variants, and the rarity of this syndrome (reported incidence of 1:10,000)^6^. The American College of Medical Genetics and Genomics (ACMG) has promulgated a framework of variant classification to guide clinician management of genetic conditions^7^. Their framework relies on functional, clinical, *in silico*, experimental/functional, and other evidence criteria to classify variants. Classifications span from benign/likely benign (B/LB) to pathogenic/likely pathogenic (P/LP) depending on various evidence strengths. Unfortunately, most variants lack sufficient clarifying information and fall as nebulous Variants of Uncertain Significance (VUS). These variants are not actionable clinically and introduce confusion and anxiety among heterozygote individuals. This is especially problematic for *RYR2* variants, given that 3912 of 7783 single nucleotide variants in the clinical genomics resource ClinVar are currently classified as VUS^8^.

Our group has recently developed an alternative approach to the ACMG system of variant interpretation, relying instead on a continuous Bayesian estimation of penetrance using a closed form beta-binomial strategy^9^. This approach addresses two key limitations of the ACMG system: 1) disease nonetheless arises in individuals harboring B/LB variants; and 2) many heterozygotes for P/LP variants will never exhibit symptoms, the problem of *incomplete penetrance*^10^. As the ACMG system, we incorporate multiple lines of evidence (structural data, *in silico*, experimental where available). Unlike the ACMG system, our approach is conditioned on prior probabilities from extensive variant curation with quantification of the estimated uncertainty. Our probabilistic approach inherently complements clinical reasoning, where diagnostic and treatment decisions are based on forecasting event probabilities (here, syncope, ventricular arrhythmias, SCD). Our group has previously deployed this strategy to study other arrhythmia syndromes including Brugada and Long QT Syndrome 1-3^9–11^. In addition to providing penetrance estimates, we have carefully curated variant *in silico*, experimental, and population evidence in a conveniently hosted ‘Variant Browser’ (variantbrowser.org) to facilitate efforts of physicians, genetic counselors, and researchers to collectively advance the care of ‘at-risk’ individuals.

Here, we apply this strategy to provide comprehensive estimates of penetrance for *RYR2*-CPVT missense variants. Our approach relies on extensive curation of the literature, population databases, and structural patterns in RyR2. We compare the value of our penetrance estimates and structural calculations to contemporary tools AlphaMissense^12^, REVEL^13^, and existing ClinVar^8^ classifications to highlight interesting insights into structural features revealed through these estimates. All data are provided at the Variant Browser (variantbrowser.org) for community uptake and use.

## Methods

### Curation of Heterozygote Variant Data

We performed a PubMed literature search with the terms ‘RYR2’ and ‘CPVT’ which resulted in 348 articles on June 29, 2022. Based on their abstracts, 179 articles underwent additional analysis. Three independent reviewers assessed each manuscript for an *RYR2* variant and affected heterozygote status, as ascertained by the authors of each paper. For individual variants, we avoided repeated heterozygote counting by excluding articles that reported duplicate patients reported by each individual institution – each study was independently cross-checked by searching for articles detailing the same variant and phenotype from the same authors/institution. We used the *RYR2* transcript ENST00000366574 throughout. We gathered unaffected *RYR2-*CPVT heterozygotes from gnomAD v.4.1.0. We have previously reported sensitivity analyses for more prevalent inherited arrhythmia syndrome genotype-phenotypes, showing that this assumption does not over inflate downstream analyses^11^. The literature curation and gnomAD dataset were summed by variant, and the count of affected and unaffected status was accordingly assigned (Supplemental Table 1; Figure 1B). Compound heterozygotes were excluded from this study.

### Penetrance Calculations

Using a previously reported strategy, we implemented Bayesian estimates of penetrance and their uncertainty using shared information across all *RYR2*-CPVT variants^10^. Formally, the observed penetrance was calculated as the fraction of affected (α) over the sum of affected (α) and unaffected (β) heterozygotes for each *RYR2* variant. The rare incidence of CPVT precluded accurate estimation due to sample size and ascertainment bias but could approximate ‘truth’ in larger datasets. For each variant i, α_,i_ and β_,i_ reflected the number of affected and unaffected heterozygous for the variant, respectively.

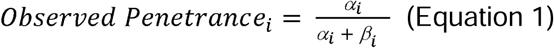

### CPVT Empirical Prior

The two parameters, α_prior, empirical_ and β_prior, empirical_, were estimated for all *RYR2* variants using a beta binomial model with weighted average (*w_i_*) (see equation 6 below) and mean squared error across all variants (implementation in GitHub). Together, the parameters defined a unique Empirical Prior applied to all *RYR2* variants. In the absence of additional variant-specific information, this value approximated the uniform penetrance of all *RYR2-*CPVT variants.

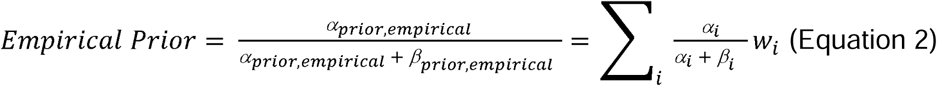

### CPVT Empirical Posterior

We refined the CPVT Empirical Prior using individual variant (i) observed affected (α_i_) and unaffected (β_i_) heterozygotes as a likelihood. This delivered a best estimate of penetrance for each variant based on observation alone.

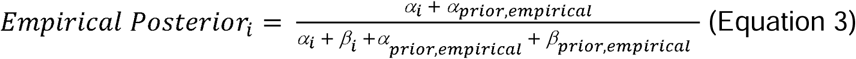

### Expectation-Maximization (EM) Algorithm

The empirical posterior enabled derivation of point estimates of a Bayesian prior and estimated uncertainty. The *RYR2*-CPVT penetrance estimates were generated using a previously described EM approach^9^. Regression used variant-specific features as independent covariates, and the empirical posteriors of those variants as dependent variables. Multiple rounds of EM were performed until iterative convergence of each probability point estimate was achieved, as informed through α_prior, empirical_ and β_prior, empirical_. This involved 3 unique steps: 1) use empirical Bayes penetrance model to derive penetrance scores; 2) penetrance estimate regression to variant features; 3) point estimate revisions to convergence.

### Bayesian Prior

Individual variant point estimates and uncertainty were defined using the new α_prior, EM,i_ and β_prior, EM,I_.

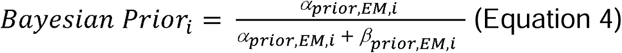

### Bayesian Posterior

If clinical data were available for the variant, then the observations (α and β in equation 1) were applied as another likelihood

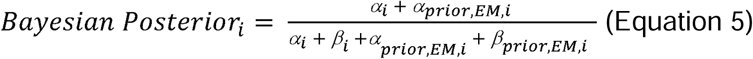

The most clinically interesting variants are ‘private mutations’. To avoid overweighting of known benign or CPVT-associated variants, as well as in penetrance estimates calculated above, we used the following weighting:

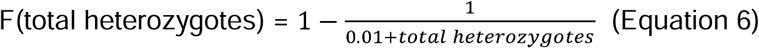

### RYR2 Variant in silico, Clinical, and Structural Features

We obtained scores for all *RYR2* missense variants from AlphaMissense^12^ and REVEL^13^, as previously described. *RYR2* variant classifications were obtained from the ClinVar resource on 5/11/2024^8^. Following current clinical practice, VUS were considered unaffected along with B/LB variants, whereas P/LP were considered affected when performing model evaluations. We analyzed the cryo-EM structure of a closed RyR2 (PDB: 7U9Q)^14^. We calculated the CPVT-density feature as a structural prediction based on the 3D-environment of each residue across RyR2, following previous implementations^15^. Regions and ‘hot-spots’ of the protein are detailed in Supplemental Table 2. When comparatively evaluating these metrics, we used a Leave One Out Cross Validation strategy to avoid over-estimation of performance. For prospective estimates, we leveraged the entirety of the available data.

### Statistical Analysis

To determine covariate performance on a binary outcome of affected/unaffected status, we used two analyses: 1) Area Under the Curve (AUC) for a Receiver-Operator Curve (ROC); and 2) AUC for a Precision-Recall Curve. These analyses were executed using R Studio Version 4, with the packages pROC and PRROC. Spearman rank order coefficients were determined in R using package wCorr. We performed 5-fold cross-validation using the Caret package in R. The dataset was randomly divided into five equal-sized folds. In each iteration, one fold was used as the test set, while the remaining four folds were used for training. This process was repeated five times, ensuring that each fold was used as a test set once. To evaluate the model’s performance, we calculated the weighted Spearman correlation between the posterior and empirical penetrance estimates. Probabilistic Brier scores were used to evaluate covariate forecasting as defined below^16^:

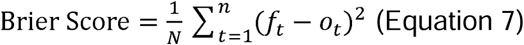

N is the quantity of observed variants, *f*_t_ is the estimated penetrance for the t^th^ variant, *o*_t_ is the observed penetrance of the t^th^ variant (Equation 3). Brier scores were calculated using different filtering allele frequences as specified in the text. All data and code necessary to perform the analyses is publicly available at https://variantbrowser.org/ and https://github.com/kroncke-lab.

## Results

### Curation of a Set of Affected/Unaffected Heterozygotes with CPVT

To develop a comprehensive set of affected and unaffected *RYR2* heterozygotes, we undertook a comprehensive literature curation of reports detailing genetic testing and clinical presentations of individuals undergoing clinical investigation (see methods; Figure 1B). An in-depth manual review of the articles identified 179 publications detailing 544 *RYR2* variants related to CPVT. Among variant heterozygotes in these reports, 1662 individuals experienced symptoms including syncope, seizures, and cardiac arrest, while 375 were asymptomatic upon extensive workup. We interrogated gnomAD for purportedly unaffected *RYR2* missense variant heterozygotes. Filtering on an allele frequency of <0.01%, we found a total of 5,159 variants harbored by 34,396 individuals. Based on previous sensitivity analyses for LQTS and the comparatively lower incidence of CPVT, we do not expect our conclusions to change based on assumption of unaffected status^9,11^.

### Results of CPVT-density and CPVT Bayesian Penetrance Calculations

Using the curated set of *RYR2* variants, we first calculated ‘CPVT-density’, a structural metric based on the 3D proximity of affected CPVT *RYR2* missense variants across the protein structure. We then determined Bayesian prior scores for variants as described in the methods (Figure 1C). These findings are hosted on the variant browser (variantbrowser.org). Figure 2A shows the distributions of observed penetrance and Bayesian penetrance. Most variants tended to have an observed penetrance of zero or one, reflecting bias from population databases as well as small numbers of heterozygotes studied in case reports. In contrast, there was a comparative ‘smoothing’ of the distribution with the Bayesian prior – no variants with estimated complete penetrance and very few with estimated complete absence of disease. Figure 2B highlights focal areas of RyR2 with enhanced penetrance. Hot-spot criteria have previously been applied in the ACMG framework to facilitate variant classification for inherited arrhythmia syndromes^17^.

**Figure 2.**
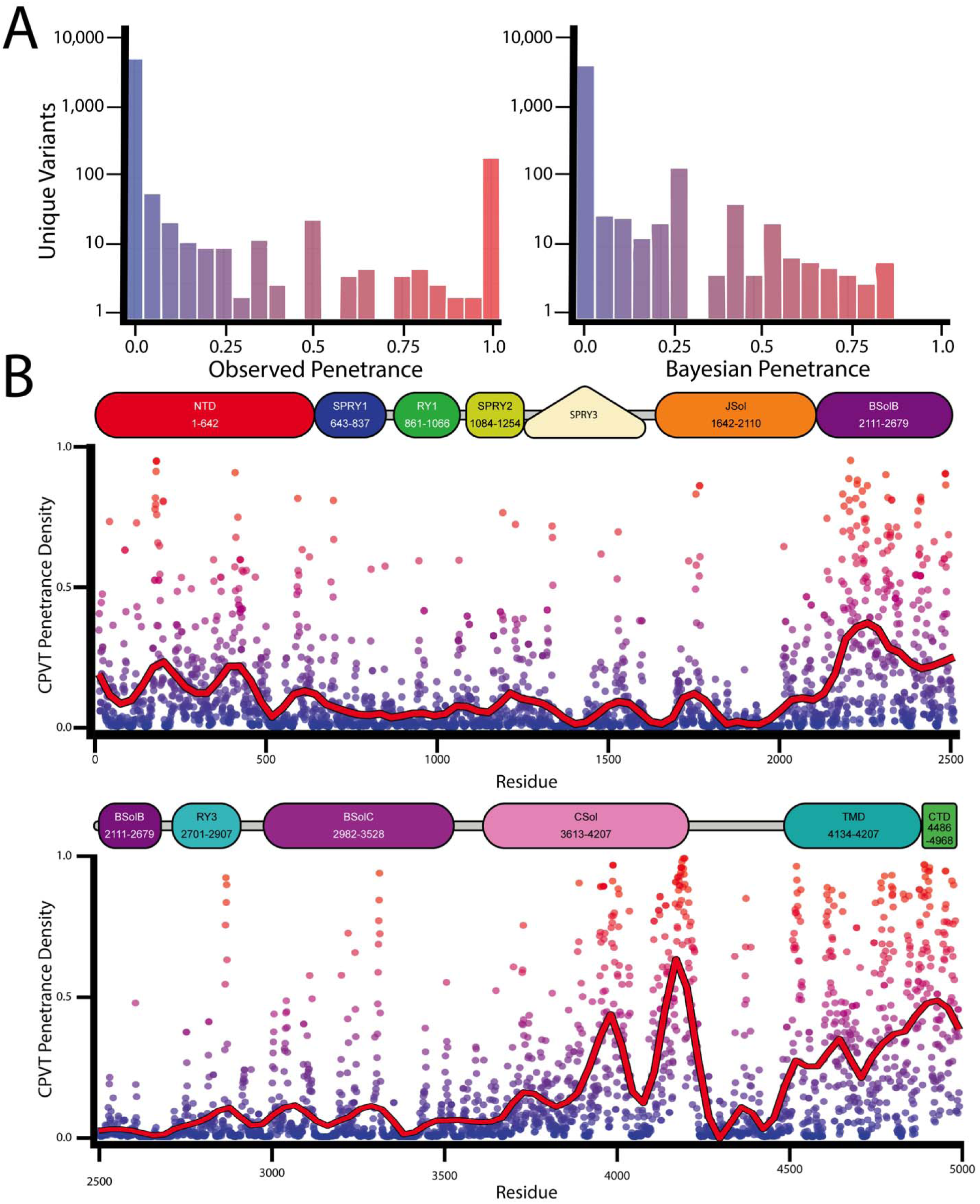
Prior Mean CPVT penetrance in high agreement with RYR2 topology. A. Histograms of *RYR2* observed and Bayesian penetrance respectively at logarithmic scale. Comparatively fewer variants have a predicted near-complete penetrance, versus the observed penetrance from limited sampling. Missense variants of *RYR2*-CPVT reported with each point. Color indicates the prior mean CPVT penetrance on a red-blue spectrum (1-0, respectively). B. Plots of residue-averaged penetrance across the length of *RYR2.* Residue numbers are reported alongside RyR2 schematic domains. Traditional ‘hot-spots’ have higher average penetrant residues (NTD, BsolB, BSolC, CSol, TMD, and CTD; 1-639, 2109-3554, 3634-4130, 4490-4886, 4887-4914, respectively).

Using a published cryo-EM structure of the closed RyR2 channel^14^, we superimposed penetrance estimates across the protein structure (Figure 3). We observed many variants with low penetrance in traditional ‘hot-spot’ domains: V186M (1/23 affected); in BsolB: R2258C (1/19 affected); in Csol: I3995V (1/8 affected); in CTD: R4959W (1/44 affected); in TMD: A4723 (1/24 affected). Interestingly, we observed moderate penetrance among canonical CPVT variants, such as R420W in the NTD (14/24 affected). Using our Bayesian penetrance observations, segments where average penetrance density exceeded >0.4 across a 100 amino acid span with more than 3 variants, only five areas met these criteria - residues 165-189 (8 variants), 401-415 (4 variants), 2082-2484 (48 variants), 3876-4204 (55 variants), 4488-4959 (54 variants; Figure 3). These data demonstrate the complexity beyond ‘hot-spot’ categorized by domain, especially when visualized on the protein structure (Figure 3).

**Figure 3.**
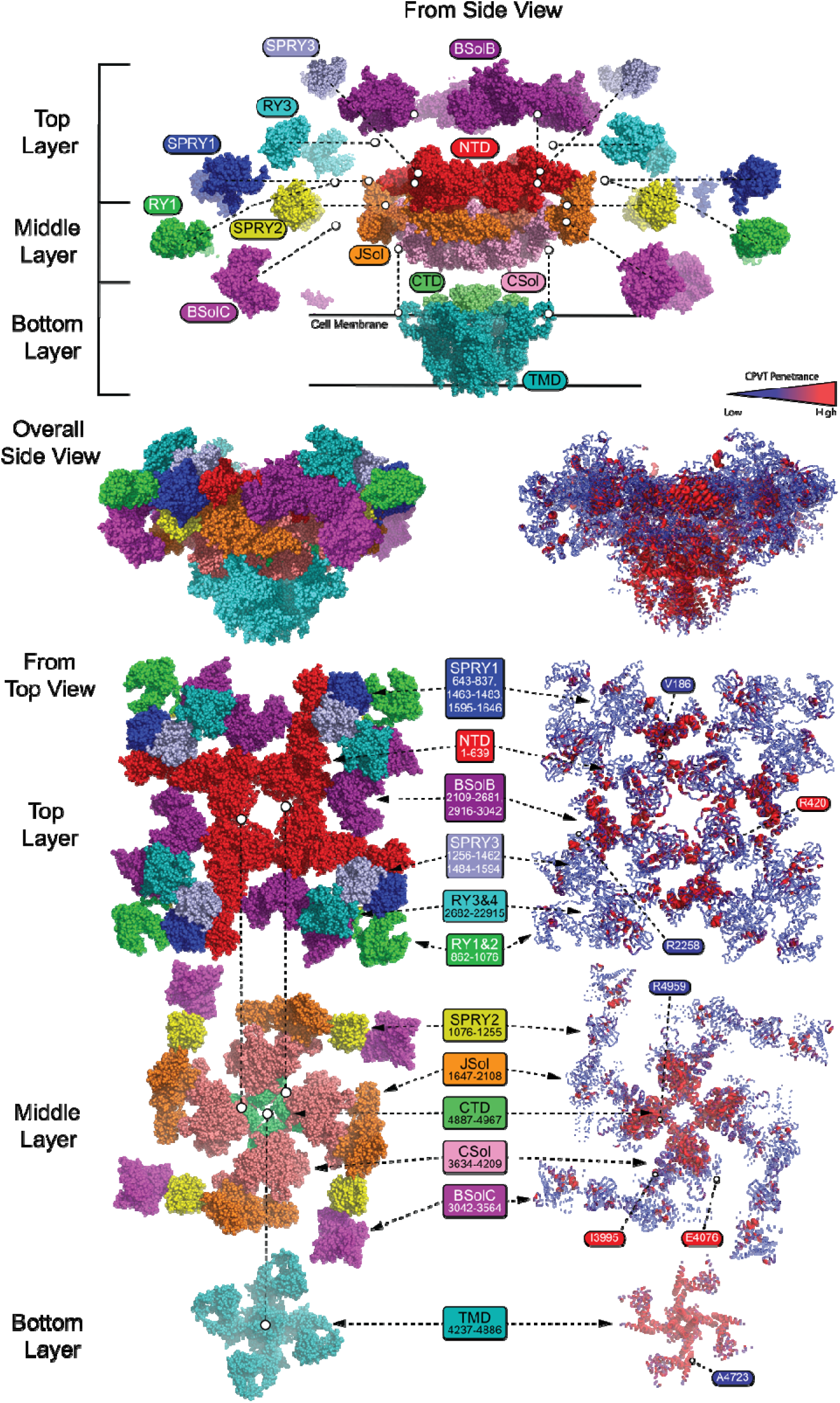
Bayesian penetrance estimates map onto structure. Cryo-EM structure of RyR2 (PDB: 7U9Q)^14^. Left depicts previously defined structural regions, right reflects penetrance estimates superimposed upon structure, averaged over each residue. Traditional ‘hot-spots’ show heterogeneity of penetrance, such as BSolB.

### Evaluation of Bayesian Penetrance, CPVT-density, in silico, and Clinical Predictors

Next, we evaluated the association of the Bayesian penetrance, CPVT-density, ClinVar, and the *in silico* predictors AlphaMissense and REVEL against the Empirical Posterior (Figure 4A; Table 1). We found that all covariates had nominally significant (p <0.05) associations. We derived the highest Spearman rank-order correlation coefficient with the Bayesian Prior (0.193), with the lowest performance by ClinVar scores (0.083). Interestingly, AlphaMissense (0.182) and REVEL (0.156) had higher coefficients than that of the CPVT density score (0.146). The above analysis is not perfectly suited as incomplete penetrance is an underlying stochastic process (a probability of disease given the presence of a variant). Accordingly, we next calculated Brier Scores for each predictor as a complementary, probabilistic evaluation metric (lower scores indicate improved performance; Table 2). We found that for Bayesian CPVT penetrance had the lowest Brier score (0.0090) compared to CPVT penetrance density (0.018), REVEL (0.018), AlphaMissense (0.018), or ClinVar (0.019).

**Figure 4.**
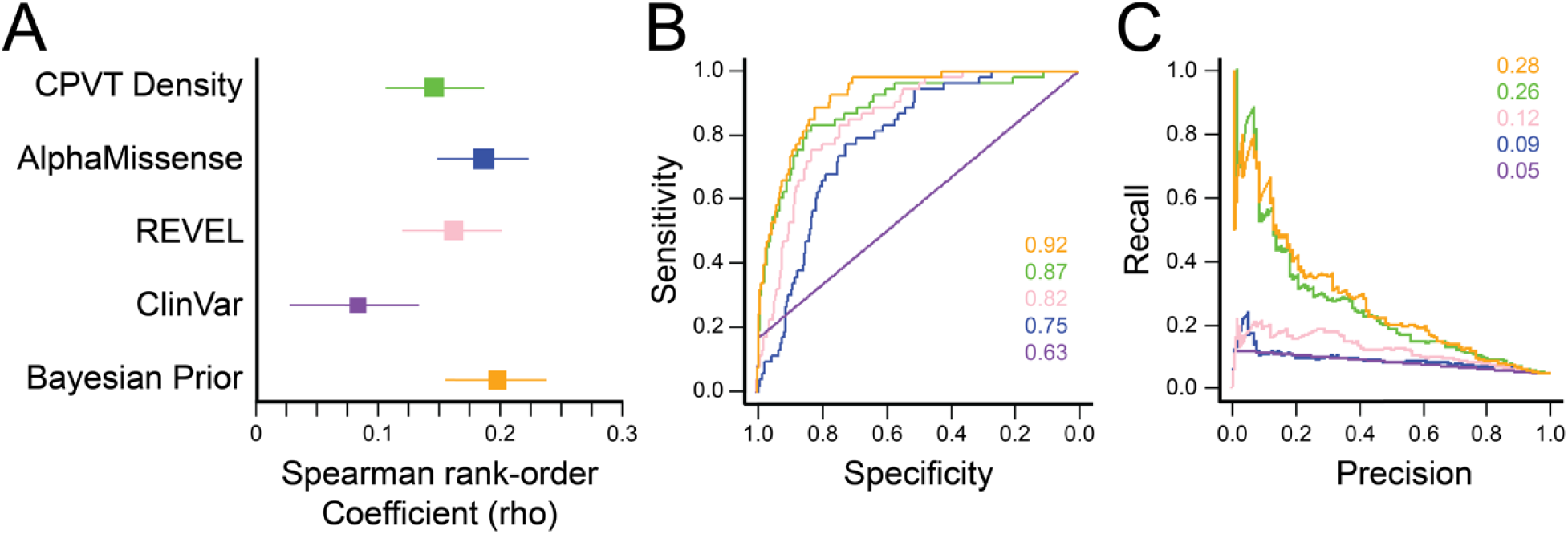
CPVT Bayesian model covariate evaluation and penetrance estimates. A. Forest Plot of Spearman correlations among model covariates. Optimal performance was obtained with Bayesian prior. B. Receiver Operating Characteristic plot of model covariates. Areas under curve displayed in inset. Colors correspond to those in Panel A. C. Precision-Recall curve of each covariate. Areas under curve displayed in inset. Colors correspond to those in Panel A. Lower score indicates better performance.

**Table 1.** Spearman correlation coefficients by CPVT model covariates.

| **Covariate** | **Allele Frequency <1%** | **All** |
| --- | --- | --- |
| Bayesian CPVT Penetrance | 0.585 | 0.589 |
| CPVT Penetrance Density | 0.146 | 0.146 |
| AlphaMissense | 0.182 | 0.186 |
| REVEL | 0.156 | 0.162 |
| ClinVar Annotation | 0.083 | 0.083 |
| Prior Mean | 0.193 | 0.198 |

**Table 2.**
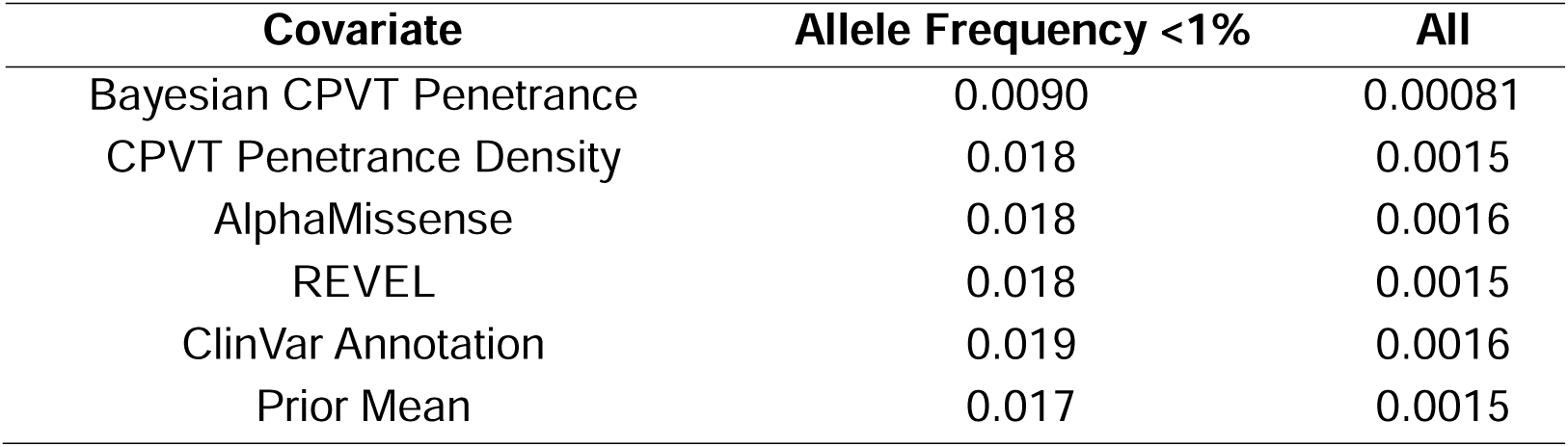
Brier Scores for Model Covariates stratified by variant frequency.

Finally, we determined Receiver Operating Characteristic curves for each model covariate (Figure 4B). The resulting areas under the curve (AUCs) from logistic models trained to predict RYR2 were as follows (AUC; [95% CI]): Revel (0.87; [0.81–0.93]), AlphaMissense (0.75; [0.68–0.82]), CPVT Penetrance Density (0.82; [0.77–0.99]), Bayes Prior (0.92; [0.84-0.99]), ClinVar (0.63 [0.51 - 0.75]). We show covariate AUCs at various cutoff thresholds in Supplemental Figure 1. As penetrance was not equally distributed across variants but more heavily weighted towards zero, we also assessed Precision-Recall AUC to address the imbalanced dataset (Figure 4C). The prior mean predictor performed well (0.28 [0.23–0.33]). CPVT-density (0.26 [0.21–0.31]) and REVEL (0.12 [0.072–0.17]) showed moderate performance, while AlphaMissense (0.090 [0.040–0.14]) and ClinVar (0.047 [−0.0030 – +0.097]) performed poorly.

## Discussion

### RYR2-CPVT Penetrance Estimates Improve Variant Interpretation

Here, we develop a predictive model of *RYR2-*CPVT missense variant penetrance to facilitate structural understanding and clinical evaluation of this lethal arrhythmia syndrome. Variant penetrance is increasingly emerging as a critical component of clinical genetics, particularly for arrhythmias and cardiomyopathies^18,19^. Whereas variant classification may help define the variant’s etiological contribution to disease (i.e. ACMG), penetrance assesses the more difficult, prospective question of whether variant heterozygotes will manifest a certadisease. Unfortunately, many studies conflate these questions, using ClinVar classifications as proxies for variant penetrance and severity. We and others have shown that ACMG variant classifications are often poor markers of disease penetrance^10,20^. Moreover, variant *prognosis* can query more granular levels of such phenotypes (age of onset, severity of manifestations, and recurrence of manifestations). Our Bayesian approach addresses both latter questions in previous work on *KCNH2*-LQT2, using a richly phenotyped clinical dataset^21^. In the present study, we are limited to penetrance assessment due to the currently less deeply phenotyped nature of our clinical datasets; however, we show that our Bayesian approach outperforms current variant effect predictors and clinical interpretations of ClinVar classifications in addressing penetrance. Our approach is fundamentally phenotype-agnostic but has been primarily deployed to study inherited arrhythmia syndromes^9–11^. Deploying our method on additional genes within cardiomyopathies and arrhythmias will complement previous disease (GENESIS)^22^ and tissue-specific (CVD-PP^23^) in this space. Moreover, the combination of close literature review, previous cohort studies, and a population database, complements recent penetrance frameworks specifically aimed at the secondary finding population^24^.

### Clinical Application of CPVT Penetrance Estimates

Genetic testing is increasingly incorporated into clinical diagnostic frameworks, including the Schwartz Score for LQTS^25^ and the Shanghai Score for Brugada Syndrome^26^. As larger genomic datasets become available, with a heightened understanding of underlying architecture, these genetic data would be expected to carry more weight. Smoothly integrating clinical observations with genetic data promises to enhance diagnosis and prognosis. In one such application, a CPVT clinical score was developed and externally validated, which insightfully combined clinical and genetic data to facilitate ‘phenotype-enhanced’ variant classification^27^. We envision that the current Bayesian penetrance estimates will provide another helpful framework for the evaluation of the genetic underpinnings a patient’s presentation.

Accordingly, the current method may facilitate the application of proactive ‘genotype-first’ approaches. As larger groups of individuals undergo whole genome sequencing, prospectively available penetrance estimates may identify individuals at risk to undergo pre-emptive clinical workup. In such a case, the prior would be based on variant data, with physical exam and diagnostic testing providing a likelihood regarding disease status. A current challenge remains the significant incomplete penetrance associated with inherited arrhythmia syndromes. For example, an early exome sequencing study quickly recognized the incomplete penetrance of reported *RYR2*-CPVT variants, which would have resulted in a prevalence of 1:150^28^. When applied correctly, this approach promises to significantly decrease morbidity and mortality by early initiation of treatment before manifestations of overt ventricular arrhythmias. Conversely, avoiding overtreatment with aggressive ICDs will decrease iatrogenic patient harm^29^. This question is especially important given that *CPVT* variants are reported as secondary findings^30^. In a recent arrhythmia-focused biobank study, the actionability of discovered *RYR2* variants was limited by the lack of P/LP annotations^31^. This challenge could be overcome by the current approach of focusing on readily available penetrance estimates. Moreover, high-throughput experimental studies of inherited arrhythmia syndrome-associated genes such as *RYR2* further promise to refine penetrance estimates and improve their validity. Our group and others have previously deployed this approach for the full arrhythmia-associated proteins *KCNH2*^21^, *KCNJ2*^32^, *KCNE1*^33^, and *CALM1-3*^34^. This confluence of advances in deeply phenotyped biobank scale cohorts, high-throughput molecular biology, and variant-based penetrance promises to facilitate the clinical implementation of personalized medicine.

## Limitations

The current Bayesian penetrance estimates are not conditioned on functional data, as high-throughput approaches to interrogate RyR2 biology are currently limited. Moreover, our clinical analyses are currently limited to diagnosis of CPVT, as we could not obtain event data at scale to further examine variant-based prognosis. We demonstrate that current predictors, including the Bayesian penetrance estimates, are unable to fully predict clinical outcomes with response to therapy.

## Conclusions

*RYR2-*CPVT Bayesian penetrance estimates provide a pathway to build upon classic variant classification. These estimates provide structural insights into regions enhanced for highly penetrant variants and will facilitate clinical genetics and interpretation of secondary findings. Variant information is conveniently hosted at the ‘Variant Browser’ for use by physicians, genetic counselors, and researchers.

## Data Availability

All data produced are available online at variantbrowser.org and https://github.com/kroncke-lab.

https://variantbrowser.org

## Ethics Declaration

All patient data was obtained retrospectively through review of previous published literature. BMK has filed the patent ‘Systems and methods for estimating variant-induced disease penetrance and estimating probability of disease occurrence based on the same; USPA 17845492’ based on previous applications of a Bayes variant penetrance estimation.

## Acknowledgements

This research was funded by the Leducq Transatlantic Network of Excellence Program 18CVD05 (BMK); and National Institutes of Health: F30 HL163923 (MJO), R01HL160863 (BMK).

## Data and Code Availability

All code used to generate the results are available at https://github.com/kroncke-lab/Bayes_Incomplete_Penetrance. All input data, CPVT-densities, and Bayes penetrance estimations are made publicly available at https://variantbrowser.org/.

