## Supplemental Information for "Structural Evaluation of *RYR2*-CPVT Missense Variants and Continuous Bayesian Estimates of their Penetrance"

**Supplementary Information**

**Supplemental Figure 1.** AUCs of each model covariate at various threshold levels.

**Supplemental Table 1.** Complete dataset of curated variants, variant-specific features, and phenotype status.

**Supplemental Table 2.** RyR2 ‘hot-spot’ region definitions.

**Supplemental Figure 1.** AUCs of each model covariate at various threshold levels.

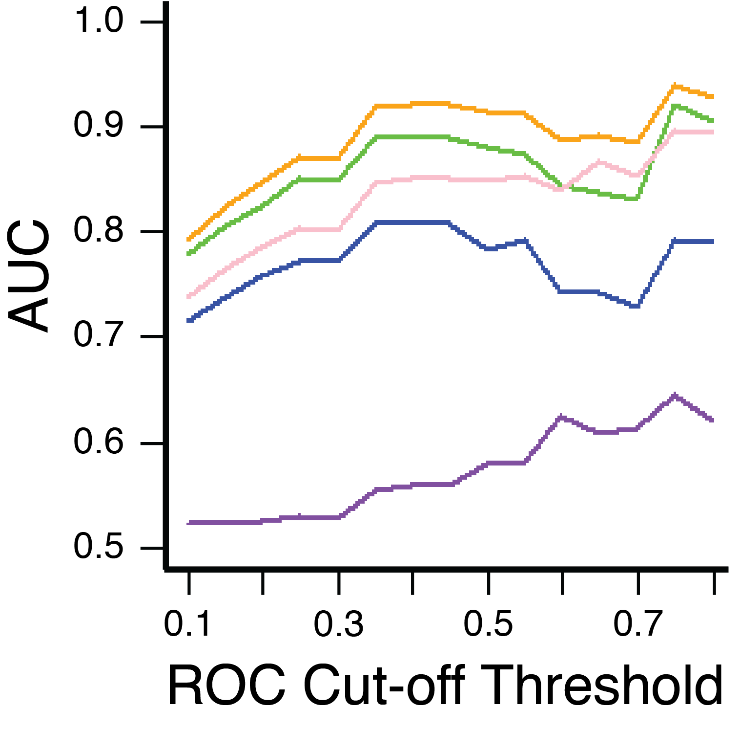

**Supplemental Table 1.** Complete dataset of curated variants, sources, and phenotype status – available as online file and at variantbrowser.org.

**Supplemental Table 2.** RyR2 ‘hot-spot’ region definitions.

| **Micro Structure** | **Symbol** | **Residue Span** |
| --- | --- | --- |
| N-terminal Domain | NTD | (1-639) |
|  | NTD-A | (1-219) |
|  | NTD-B | (220-408) |
|  | NSol | (409-639) |
| SPRY Domain |  | (640-1646) |
|  | SPRY1 | (640-861,1463-1483,1595-1646) |
|  | SPRY2 | (1076-1255) |
|  | SPRY3 | (1256-1462,1484-1594) |
|  | RY1&2 | (862-1076) |
| Junctional solenoid | Jsol | (1647-2108) |
| Bridging solenoid | BSol | (2109-3564) |
|  | BSol1 | (2109-2681,2916-3042) |
|  | BSol2 | (3042-3344) |
|  | BSol3 | (3345-3564) |
|  | RY3&4 | (2682-2915) |
| Shell-core linker peptide | SCLP | (3565-3633) |
| Core solenoid | CSol | (3634-4130) |
|  | EF1&2 | (4016-4090) |
| Thumb and forefingers domain | TaF | (4131-4209) |
| Transmembrane domain | TM | (4237-4886) |
|  | Sx | (4237-4310) |
|  | pVSD | (4480-4750) |
|  | Pore | (4751-4886) |
| C-terminal domain | CTD | (4887-4967) |
|  | ZnF | (4887-4914) |
